# Characterizing cerebral metabolite profiles in anorexia and bulimia nervosa and their associations with habitual behavior

**DOI:** 10.1101/2021.09.12.21263466

**Authors:** Margaret L. Westwater, Alexander G. Murley, Kelly M.J. Diederen, T. Adrian Carpenter, Hisham Ziauddeen, Paul C. Fletcher

**Affiliations:** Department of Psychiatry, University of Cambridge, Herchel Smith Building, Addenbrooke’s Hospital, Cambridge CB2 0SZ, UK; Department of Radiology and Biomedical Imaging, Yale University School of Medicine, New Haven, CT, USA; Department of Clinical Neurosciences, University of Cambridge, Herchel Smith Building, Addenbrooke’s Hospital, Cambridge CB2 0SZ, UK; Institute of Psychiatry, Psychology and Neuroscience, King’s College London, London WC2R 2LS, UK; Wellcome Trust MRC Institute of Metabolic Science, University of Cambridge, Cambridge Biomedical Campus, Cambridge CB2 0QQ, UK

**Author notes:** Please address correspondence regarding this manuscript to: Margaret L. Westwater, Douglas House, 18b Trumpington Road, Cambridge, CB2 8AH, United Kingdom.

**Keywords:** Eating disorders, inferior frontal cortex, goal-directed behavior, magnetic resonance spectroscopy

## Abstract

**Background:** Anorexia nervosa (AN) and bulimia nervosa (BN) are associated with altered brain structure and function, as well as increased habitual behavior. This neurobehavioral profile may implicate neurochemical changes in the pathogenesis of these illnesses. Altered glutamate, *myo*-inositol and *N-*acetyl aspartate (NAA) concentrations are reported in restrictive AN, yet whether these extend to binge-eating disorders, or relate to habitual traits in affected individuals, remains unknown.

**Methods:** Using single-voxel proton magnetic resonance spectroscopy, we measured glutamate, *myo*-inositol and NAA in 85 women [n=22 AN (binge-eating/purging subtype; AN-BP), n=33 BN, n=30 controls]. Spectra were acquired from the right inferior lateral prefrontal cortex and the right occipital cortex. To index habitual behavior, participants performed an instrumental learning task and completed the Creature of Habit Scale. Exploratory analyses examined associations between metabolites and habitual behavior.

**Results:** Women with AN-BP, but not BN, had reduced myo-inositol and NAA concentrations relative to controls in both voxels. Patient groups had intact performance on the instrumental learning task; however, both groups reported increased routine behaviors compared to controls. Women with BN also reported greater automatic behaviors, and automaticity was related to reduced prefrontal glutamate and NAA in the AN-BP group.

**Discussion:** Findings extend previous reports of reduced *myo*-inositol and NAA levels in AN to AN-BP, which may reflect disrupted axonal-glial signaling. Although we found inconsistent support for increased habitual behavior in AN-BP and BN, we identified preliminary associations between prefrontal metabolites and automaticity in AN-BP. These results provide further evidence of unique neurobiological profiles across binge-eating disorders.

## 1. Introduction

Bulimia nervosa (BN) and the binge-eating and purging diagnostic subtype of anorexia nervosa (AN-BP) are complex psychiatric conditions that share core symptoms, including recurrent binge-eating and compensatory behaviors (e.g., vomiting). The primary diagnostic distinction between AN-BP and BN relates to an individual’s body mass index (BMI), wherein individuals with AN-BP are underweight and those with BN are not (1). Neuroimaging studies have begun to characterize the neural correlates of these illnesses, identifying alterations in the structure and function of brain regions that subserve learning, self-regulatory control and bodily perception in affected individuals (2–6). However, as perturbed metabolic functioning is increasingly implicated in AN and, to a lesser degree, BN (7, 8), a more comprehensive understanding of their neurocognitive mechanisms will require consideration of interacting metabolic processes, both peripherally and within the brain.

Proton magnetic resonance spectroscopy (^1^H-MRS) enables *in vivo* measurement of tissue metabolite concentrations, which can lend insight into various physiological processes within a given anatomical region (9). In the context of mental illness, selecting the region based on its putative relevance to the disorder under examination can offer useful ways to begin to understand structure-function relations and their disruption. Several studies have assessed cerebral metabolites in eating disorders using ^1^H-MRS, yet these have been primarily limited to AN, where few consistent findings have emerged. Schlemmer et al. (10) reported increases in the ratio of choline (a marker of cell membrane integrity) to total creatine (a stable marker of energy metabolism), in parieto-occipital white matter (WM) among adolescents with AN. Affected adolescents also demonstrated reduced relative concentrations of *N*-acetyl aspartate (NAA) in parieto-occipital WM, and NAA reductions have been observed in the anterior insula in adult women with AN (11). Although NAA is highly concentrated in grey matter (GM) and often considered a marker of neuronal density (12), it has various functional roles, including osmoregulation, myelin synthesis, neuron-glia signaling and glutamate turnover [reviewed by (13)]. Moreover, adults with AN have reduced creatine and *myo-*inositol [a marker of glial cell integrity (14)] concentrations in dorsolateral prefrontal cortex GM, and lower levels of the composite metabolite ‘Glx’, which reflects both glutamate and glutamine, in the anterior cingulate cortex [ACC; (15)]. Findings of reduced glutamate and *myo*-inositol have been replicated in both adolescent and adult AN across various brain regions, including medial frontal, anterior cingulate and occipital cortices and the putamen (16, 17).

Neural biochemistry has yet to be explicitly studied in BN, but previous examination of mixed samples of patients with AN and BN lends some insight into potential metabolic alterations in this illness. Emerging adults with either AN or BN have reduced lipid concentrations in frontal and occipital WM, as well as decreased *myo*-inositol in frontal WM (18). However, a relatively small sample of adults with AN or BN did not differ significantly from matched controls in spectra acquired from the ACC (19).

Such observations are potentially informative since altered neural chemistry in eating disorders could suggest a role for these metabolites in higher-order cognition (15), such as instrumental learning. The persistent and often intractable nature of binge-eating and compensatory episodes could suggest a role for dysregulated learning in the pathogenesis of AN-BP and BN, where biases toward stimulus-response (‘habitual’) rather than action-outcome (‘goal-directed’) behavior may explain, in part, recurrent loss-of-control eating (20–22). This distinction can also be framed in terms of model-free and model-based processes, respectively (23, 24). Although both goal-directed and habitual systems support instrumental learning, these systems have largely dissociable neural circuits. Whereas dorsomedial striatal and ventromedial prefrontal areas facilitate goal-directed learning and action (25), corticostriatal loops encompassing the posterior putamen and premotor cortex are traditionally associated with habitual responses (26). Reduced glutamate turnover in the putamen has been linked to greater habitual tendencies in patients with cocaine use disorder (27); however, it remains unknown if such an association would generalize to other regions implicated in habitual control. For example, the inferior lateral prefrontal cortex (ilPFC) has been shown to track both model-free (i.e., habitual) and model-based (goal-directed) systems, thus determining the relative control of each system over behavior (28). Moreover, theta-burst stimulation of the right ilPFC attenuates goal-directed and increases habitual behavioral control (29), supporting a causal role for this region in achieving an optimal balance between goal-directed and habitual processing. Assessment of right ilPFC metabolite profiles could therefore lend complementary insight into the neurobiological mechanisms that guide goal-directed and habitual responding.

Here, we sought to extend previous reports of altered cerebral metabolites and greater habitual tendencies in women with eating disorders by examining these processes, for the first time, in women with AN-BP, BN and matched controls. Participants underwent ^1^H-MRS scanning and behavioral testing within an inpatient study session. On the basis of prior literature (11, 15–18), glutamate, *myo*-inositol and NAA levels were measured in the right ilPFC and the right occipital cortex, which served as a control region. To relate observations at the neurometabolic level to goal-directed and habitual systems, participants performed an instrumental learning task, which manipulated stimulus-outcome conflict to distinguish between goal-directed and habitual learning (25, 30, 31). Participants also completed the Creature of Habit Scale [COHS; (32)], a validated measure of routine and automatic responses in daily life. As laboratory paradigms assess experimentally induced habits that are substantially less practiced than those in an individual’s day-to-day life, the COHS serves as a complementary index of the habitual system. We anticipated reduced glutamate, *myo*-inositol and NAA concentrations in women with AN-BP and BN compared to control participants. Additionally, we predicted that both patient groups would show a shift in balance toward habitual responding, as evidenced by relative insensitivity to outcome devaluation after learning a set of stimulus-outcome pairings and greater COHS scores. Finally, exploratory analyses examined whether ilPFC glutamate, *myo*-inositol and NAA levels were associated with a shift toward habit-based responding.

## 1. Methods and Materials

### 2.1 Participants

Eighty-five women (M±SD = 23.96±3.98 years) were recruited via posted advertisements and eating disorder services in the Cambridgeshire, UK area to three participant groups: AN-BP (n=22), BN (n=33) and matched control participants (n=30). Patient participants were free of alcohol or substance use disorders within the past 6 months, and they had no lifetime history of serious mental illness (e.g., bipolar disorder, schizophrenia) or neurodevelopmental disorders. Control participants reported no current or lifetime psychopathology. Exclusion criteria for all participants included: left handedness, MRI contraindications, estimated IQ<80, BMI>29.9 kg/m^2^, neurological or cardiovascular disease, lactation, prior bariatric surgery and high nicotine dependence (indexed via the Fagerstrom Test for Nicotine Dependence (33)). All participants provided written, informed consent prior to undergoing any study procedures. The Cambridge East Research Ethics Committee approved the study (Ref. 17/EE/0304), and all procedures were performed in accordance with local regulations.

### 2.2 Study procedures

As a detailed explanation of the study protocol has been reported previously (2, 34), we provide a summary here. Following a telephone screening, potential volunteers completed an outpatient screening session prior to scanning, which included body composition testing, blood sampling, cognitive testing and two, semi-structured clinical interviews [e.g., the SCID-5 (35) and EDE v16 (36)] to establish current diagnoses of AN-BP, BN and comorbid psychiatric conditions. Eligible participants (n=85) then attended a two-day, inpatient study session, which included a cognitive testing and questionnaire battery, repeated MRI scanning and serial blood sampling to enable measurement of circulating hormones under neutral and stressful conditions. On each day of the inpatient session, participants underwent either an acute stress induction or control task during MRI scanning. All participants underwent ^1^H-MRS scanning on their ‘control’ day, and spectra were acquired prior to completion of both the fMRI task and induction. During spectra acquisition, participants were instructed to remain still, keep their eyes open and gaze at a fixation cross. The order of acquisition from each voxel of interest (VOI) was randomized across participants.

### 2.3 Slips of action paradigm

Participants performed a four-stage instrumental learning task, which had been adapted from prior work to include photos of animals, as opposed to fruit, as training and outcome stimuli [Figure 1; (30, 31)]. Throughout the discrimination training phase, participants were required to learn which response (“M” or “Z” on a standard keyboard) to a given stimulus lead to a valued outcome (i.e., one that was worth points). Participants received trial-by-trial feedback over the course of 8 blocks (12 trials each), and they were told that the aim of the task was to earn as many points as possible. The second stage of the task tested participants on their outcome-action knowledge. In this stage, each trial (36 total) presented two of the outcomes from the discrimination training, where one outcome was devalued (indicated by a superimposed red “X”) and not worth any points. Participants were instructed to make a response that previously led to the valued outcome; however, no feedback was provided. The third and fourth stages of the task tested habits (via the slips-of-action test) and general inhibitory control (via a control test), and the order of these stages was randomized across participants. For the slips-of-action test, participants were shown all of the outcomes from the initial training, and two outcomes had a red “X” overlaid, indicating that they were no longer worth points. Participants were then shown the stimuli from the discrimination training, and they were told to only respond to stimuli that were associated with valued outcomes and withhold responses to stimuli associated with devalued outcomes. As such, responses to stimuli with devalued outcomes are considered ‘slips-of-action’ that indicate an overreliance on stimulus-response learning. Finally, participants performed the control test, which had an identical structure to the slips-of-action test with one key distinction: for this test, the discriminative stimuli were devalued (with a red “X”) instead of the outcomes. Stimuli were presented in rapid succession, and participants were told to respond to ‘valued’ stimuli and withhold responses for ‘devalued’ stimuli. For all stages of the task, stimuli were presented for a maximum of 2000ms with a 1500ms interstimulus interval. Following the task, participants completed a brief questionnaire about their knowledge of the stimulus, outcome and response associations (see Supplementary Material).

**Figure 1.**
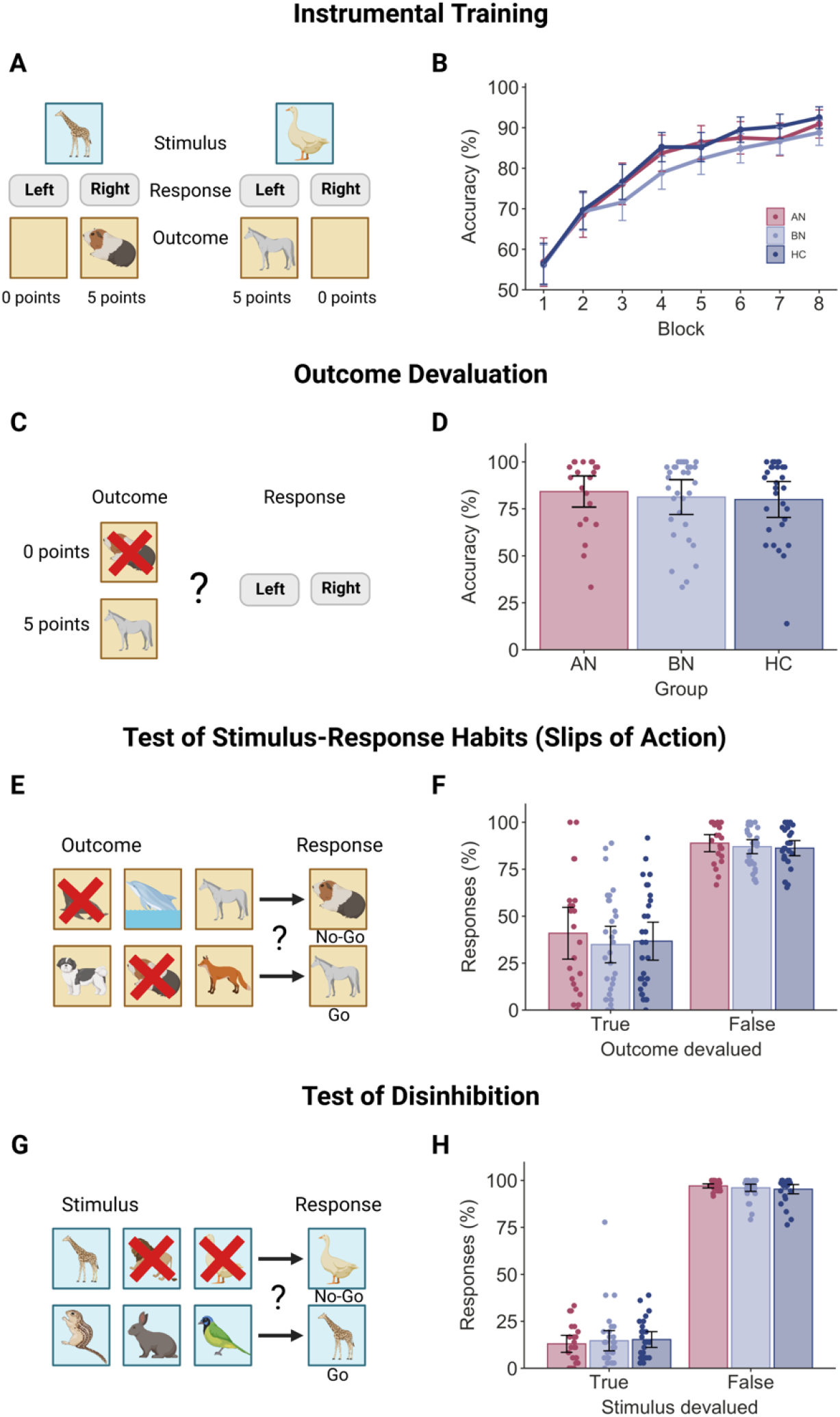
Performance on the Slips of Action task. **A)** During the instrumental training stage, participants learned stimulus-action-outcome contingencies by trial and error. Trial-by-trial feedback consisted of either a picture of another animal and the number of points gained, or an empty box and no points gained. **B)** Discrimination learning improved across each 12-trial block (p<.0001), and accuracy did not differ significantly between groups (all p’s >.05). **C)** For the second stage, participants were presented with two outcomes simultaneously, where one outcome was devalued (as indicated by a superimposed ‘X’) and no longer worth points. **D)** Patient groups had nonsignificant differences in accuracy (all p’s >.05), which would suggest intact sensitivity to outcome devaluation in both AN-BP and BN groups. **E)** Next, participants performed the ‘Slips of Action’ test. At the start of each block, participants were shown all six outcome animals, and two were devalued. Participants then selectively responded to discriminative stimuli that were associated with a reward and withheld responses to devalued stimuli. **F)** Participants made significantly more responses to valued outcomes (p<.0001), and a group-by-stimuli interaction was nonsignificant. **G)** The Test of Disinhibition served as a control task by indexing participants’ inhibitory control: participants were shown the six discriminative stimuli, and two animals were devalued. Participants were instructed to respond only to valued stimuli. **H)** Participants’ response rate was significantly higher for valued stimuli (p<.0001), and response rate did not differ between groups. The order of Stages 3 and 4 (i.e., the Slips of Action and Test of Disinhibition stages) was counterbalanced across participants. Error bars = 95% CI. Created with BioRender.com.

### 2.4 MRI data acquisition

Participants underwent MRI scanning on a 3T Siemens Skyra^Fit^ scanner (Erlangen, Germany) with a 32-channel, GRAPPA parallel-imaging head coil. T1-weighted anatomical images (TE=2.95ms, TR=2300ms, flip angle=9°, acquisition matrix=256×256mm, 1.0mm isotropic resolution) were acquired for the placement of two, 20mm isotropic VOIs in the right ilPFC and right medial occipital cortex (Figure 2A&B).

**Figure 2.**
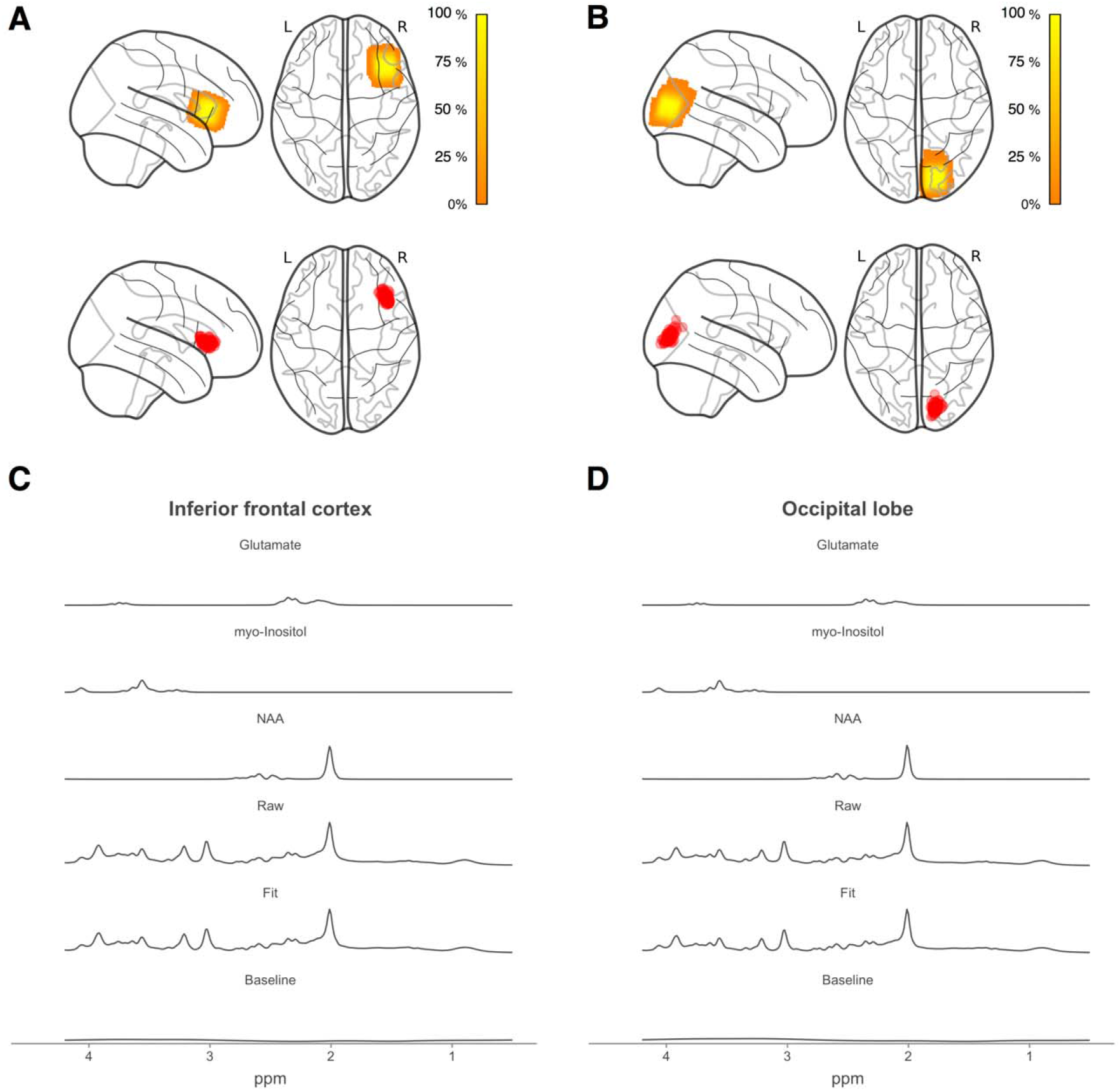
Spectroscopy voxel location and composition. Voxel density maps of **(A** inferior frontal (MNI_X,Y,Z_ = 40, 21, 9) and **(B)** occipital (MNI_X,Y,Z_ = 17, -87, 14) voxel placement overlaid on a glass brain template. MNI coordinates refer to center of mass of the group composite voxel. Color bars indicate percentage overlap across all participants. Centroids for each participant’s ^1^H-MRS voxel are shown in red. Mean spectra from all participants for **(C)** inferior frontal and **(D)** occipital voxels, showing the raw data, LCModel fit, baseline, glutamate, *myo*-inositol and *N*-acetyl aspartate fits. NAA = *N*-acetyl aspartate. Code for voxel density and centroid figures (72, 73) was retrieved from: https://github.com/nwd2918/MRS-voxel-plot.

For each VOI, first- and second-order B_0_ field shims were adjusted using 3D gradient-echo shimming. RF pulses were then calibrated in semi-LASER (sLASER) and for water suppression by 1) monitoring the water signal intensity across increasing levels of RF power and 2) choosing the settings for maximum signal (37). Spectra were acquired using semiLASER sequence (total TE=28ms, TR=5000ms, 64 transients), which uses a single slice-selective 90° excitation pulse (38–40). Water signal suppression was achieved using variable pulse power and optimisation relaxation delays (VAPOR) water suppression, with one additional pulse following the seventh VAPOR pulse to optimally reduce in vivo water signal (41). VAPOR pulses were interleaved with outer volume suppression pulses to control for saturation effects. In addition to metabolite spectra, unsuppressed water reference scans were collected used to remove residual eddy current effects.

### 2.5 Analytic plan

#### 2.5.3 Data analysis - Behavior

Linear mixed-effects models [LMMs; R package ‘nlme’ (42)] evaluated group differences in performance [accuracy and reaction time (RT)] during the instrumental learning, test of disinhibition and slips-of-action stages of the instrumental learning task, and we tested group differences in sensitivity to outcome devaluation (stage 2) using multiple regression (see Supplementary Material).

Two participants failed to perform above chance level (i.e., >50% accuracy) on the final block of the instrumental learning stage. Since, in the absence of learning, performance on the slips of action task becomes uninterpretable, these participants were excluded from further behavioral analyses. One additional participant was excluded from analysis of the test of disinhibition due to a technical error.

#### 2.5.2 Data analysis - Neuroimaging

Pre-processing of ^1^H-MRS was completed in Matlab (v2016a; The Mathworks, Natick, MA, USA), using the MRSpa package (https://www.cmrr.umn.edu/downloads/mrspa/; see Supplementary Material for details). We quantified the sLASER spectra using LCModel [v6.3.3; (43, 44); Figure 2C&D] in line with consensus recommendations for clinical MRS research (45). Metabolite concentrations were estimated with water scaling, using a basis set comprised of metabolites between 0.5 and 4.2 part per million (including glutamate, *myo*-inositol, and NAA; see Table 2 for tissue-corrected concentrations). Only metabolites that were quantified with Cramer-Rao lower bounds (CRLB) values <30% were classified as detected. CRLB values >20% were classified as outliers; however, no participants were excluded on this basis (see Supplementary Table 1).

**Table 1.** Sample demographic and clinical characteristics

| Characteristic | AN (*n* = 22) |  | BN (*n* = 33) |  | HC (*n* = 30) |  | Analysis |  |
| --- | --- | --- | --- | --- | --- | --- | --- | --- |
| | M | SD | M | SD | M | SD | $\chi^2$(df), *F*(df),<br>W, *t*(df) | *p* |
| Age (y) | 24.6 | 4.7 | 23.6 | 3.9 | 23.9 | 3.5 | $\chi^2$(2)= 0.8 | .69 |
| BMI (kg/m<sup>2</sup>) | 16.4 | 1.4 | 22.0 | 2.4 | 21.9 | 2.1 | $\chi^2$(2)= 48.4 | <.001 |
| IQ <sup>a</sup> | 116 | 5 | 114 | 5 | 114 | 5 | $\chi^2$(2)= 3.2 | .21 |
| Age of onset (y) | 15.6 | 2.4 | 16.2 | 3.1 | - | - | *t*(51.8) = -0.8 | .42 |
| Illness duration (y) | 9.0 | 5.8 | 7.4 | 4.0 | - | - | *t*(34.4) = 1.1 | .27 |
| Beck Depression Inventory | 35.3 | 12.0 | 32.7 | 10.5 | 2.4 | 2.8 | $\chi^2$(2)= 57.7 | <.001 |
| Trait Anxiety Inventory | 63.1 | 10.4 | 62.8 | 7.3 | 33.0 | 6.9 | *F*(2) = 151.1 | <.001 |
| Eating Disorder Examination Questionnaire | 4.4 | 0.8 | 4.6 | 0.8 | 0.2 | 0.2 | $\chi^2$(2)= 58.0 | <.001 |
| Eating Disorder Examination ratings <sup>b</sup> |  |  |  |  |  |  |  |  |
| Objective binge-eating episodes | 38.1 | 47.9 | 23.0 | 29.1 | - | - | W = 317.5 | .43 |
| Subjective binge-eating episodes | 9.5 | 12.8 | 6.6 | 6.2 | - | - | W = 341.5 | .93 |
| Vomiting episodes | 43.5 | 51.6 | 24.2 | 31.0 | - | - | W = 304.0 | .31 |
| Laxative episodes | 1.1 | 3.4 | 2.0 | 3.9 | - | - | W = 421.5 | .18 |
| Exercise episodes | 7.4 | 13.6 | 10.9 | 9.4 | - | - | W = 478.5 | .04 |
| | N | % | N | % | N | % | $\chi^2$(df) | *p* |
| Comorbid diagnoses |  |  |  |  |  |  |  |  |
| Anxiety disorder | 3 | 13.6 | 3 | 9.1 | - | - | $\chi^2$(1)= 0.3 | .69 |
| Major depressive episode | 15 | 68.2 | 16 | 48.5 | - | - | $\chi^2$(1)= 2.1 | .15 |
| Personality disorder | 2 | 9.1 | 5 | 15.2 | - | - | $\chi^2$(1)= 0.4 | .69 |
| Any current treatment | 13 | 59.0 | 15 | 45.5 | - | - | $\chi^2$(1)= 1.0 | .32 |
| Psychotherapy | 9 | 40.9 | 9 | 27.3 | - | - | $\chi^2$(1)= 1.1 | .29 |
| Medication | 10 | 45.5 | 10 | 30.3 | - | - | $\chi^2$(1)= 1.3 | .25 |
| Prior restrictive AN | 14 | 63.6 | 10 | 30.3 | - | - | $\chi^2$(1)= 6.0 | .01 |

**Note:** <sup>a</sup>Estimated full-scale IQ from the National Adult Reading Test. <sup>b</sup>EDE ratings reflect counts over the previous 28 days. Group differences were evaluated using one-

**Table 2.**
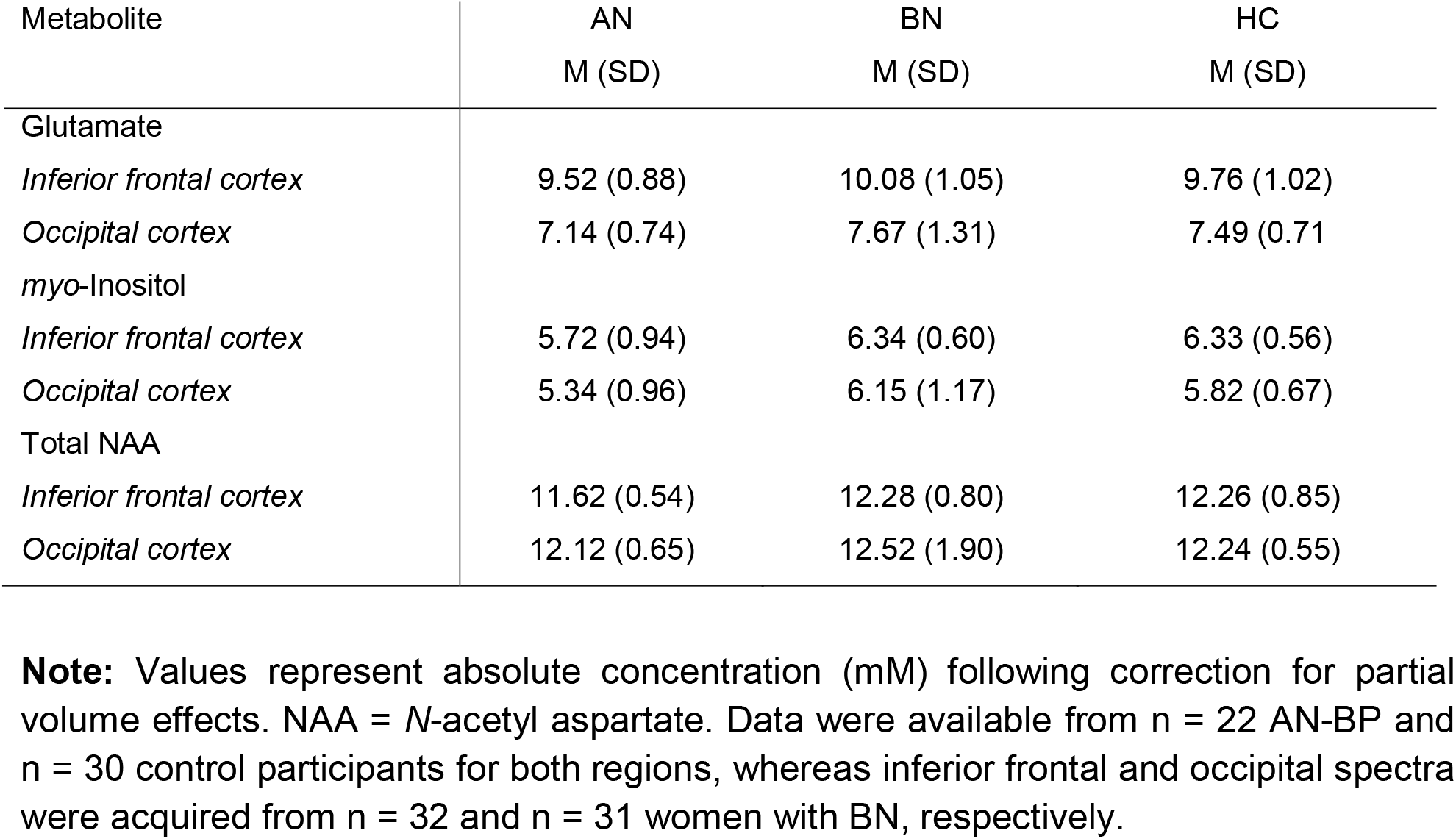
LCModel corrected metabolite concentrations by group and region

As estimation of ^1^H-MRS metabolite concentrations varies across tissue types and CSF (46, 47), we calculated the fraction of GM, WM and CSF within each VOI (see Supplementary Material). The mean percentage of each tissue was entered into the following equation from Egerton et al. (48) to generate corrected metabolite concentrations (*Mcorr*) of metabolite *M*:

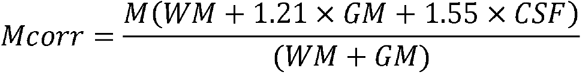

The proportion of GM, WM and CSF in each VOI did not differ significantly by group (all p’s>.05); however, LMMs indicated reduced proportions of CSF (β(SE)=-3.76(0.26), t(80)=-14.50, p<.001) and GM (β(SE)=-12.58(0.82), t(80)=-15.37, p<.001) in occipital VOIs. These reductions were offset by increases in the proportion of WM in occipital relative to right IFG VOIs (β(SE)=16.77(0.97), t(80)=17.24, p<.001).

In keeping with behavioral analyses, we examined group and regional differences in the three metabolites of interest, glutamate, *myo*-inositol, and NAA, using LMMs. Fixed effects of group (AN-BP>HC, BN>HC) and region (ilPFC>occipital) were entered into each model, and random intercepts for region were nested within the subject’s random effect. To account for multiple models, we applied a Bonferroni correction for 3 tests, yielding an alpha threshold of .05/3=.017. Results with corresponding p-values >.017 but <=.05 are referred to as ‘nominally significant.’ Visual inspection of quantile-quantile plots was used to determine the normality of model residuals. Given the potential confounding effects of psychotropic medication on metabolite concentrations (49), we conducted sensitivity analyses controlling for current medication use.

Finally, exploratory multiple regression analyses examined associations between metabolite levels and habitual responding and whether these were specific to the ilPFC. Specifically, we tested whether main and interaction effects of group, ilPFC and occipital metabolite levels were associated with COHS subscales and the mean difference score of responses during the slips-of-action test. Exploratory results were considered statistically significant at alpha=.05.

One participant was excluded, and subsequently offered clinical follow-up, on the basis of WM abnormalities, and spectra from the occipital VOI could not be acquired for another participant due to a technical error. In total, ilPFC and occipital spectra from 84 and 83 participants, respectively, were analyzed via LCModel. Finally, we classified outlying corrected concentration values as those ±3SD from the mean, which resulted in the exclusion of one observation for *myo*-inositol and NAA models, respectively.

## 2. Results

### 3.1 Behavioral results

#### Instrumental learning

During the initial training, a significant effect of block on accuracy (β(SE)=4.44(0.21), t=21.03, p<.0001) and RT (β(SE)=-0.13, t=-17.50, p<.0001) indicated that participants learned to select the response that yielded a rewarding outcome for each stimulus (see Figure 1A). Neither accuracy nor RT differed between groups, and group-by-block interaction effects were nonsignificant (all p’s>.05).

#### Sensitivity to outcome devaluation

Neither outcome-action knowledge (F(2,78)=0.26, p=.77; see Figure 1C) nor RT (F(2,78)=0.30, p=.74) differed significantly between groups.

#### Test of disinhibition

Participants responded more frequently to valued stimuli relative to devalued stimuli on the baseline discrimination task (β(SE)=81.67(1.49), t=54.97, p<.0001; Figure 1E). However, both the main effect of group and a group-by-stimulus-value interaction effect on accuracy were nonsignificant, indicating comparable performance across groups (all p’s >.05). All main and interaction effects on RT were nonsignificant.

#### Slips of action

The slips-of-action test required participants to selectively respond to stimuli that were associated with valuable outcomes and withhold responses to those with devalued outcomes. All participants responded more frequently to valued outcomes (β(SE)=50.12(3.26), t=15.38, p<.0001; Figure 1G), and a nonsignificant group-by-outcome-value interaction indicated intact outcome sensitivity in both patient groups (p=.87). RTs did not differ between groups or by outcome value (all p’s>.05). Moreover, the mean difference in responding to valued versus devalued outcomes—a measure of relative goal-directed and habitual control—did not differ significantly between groups (F(2,80)=0.33, p=0.72). This would suggest that women with AN-BP, BN and control participants were similarly sensitive to devalued outcomes.

### 3.2 Neuroimaging results

*Myo*-inositol levels were significantly reduced in AN-BP, but not BN, relative to healthy women (β(SE)=-0.55(0.18), t(81)=-2.98, p=.004; Figure 3A). Although *myo*-inositol levels were greater in the ilPFC relative to occipital cortex (β(SE)= 0.42(0.06), t(81)=7.13, p<.0001), regional concentration did not differ significantly between groups (all p’s >.05). Similarly, NAA was reduced in AN-BP compared to control participants (β(SE)=-0.42(0.15), t(81)=-2.75, p=.007) and increased in the ilPFC relative to occipital VOI (β(SE)=0.55(0.10), t(81)=5.60, p<.0001). Nonsignificant differences were observed in BN (p=.60), and a group-by-VOI interaction was nonsignificant (all p’s >.05). Diagnostic status was not significantly related to glutamate concentration; however, as with the other metabolites, glutamate levels were increased in prefrontal cortex (β(SE)=2.36(0.15), t(82)=15.69, p<.0001). There were no significant group differences in MRS spectra quality metrics (Supplementary Table 1), which suggests that results were not confounded by group differences in, e.g., head motion artifacts.

**Figure 3.**
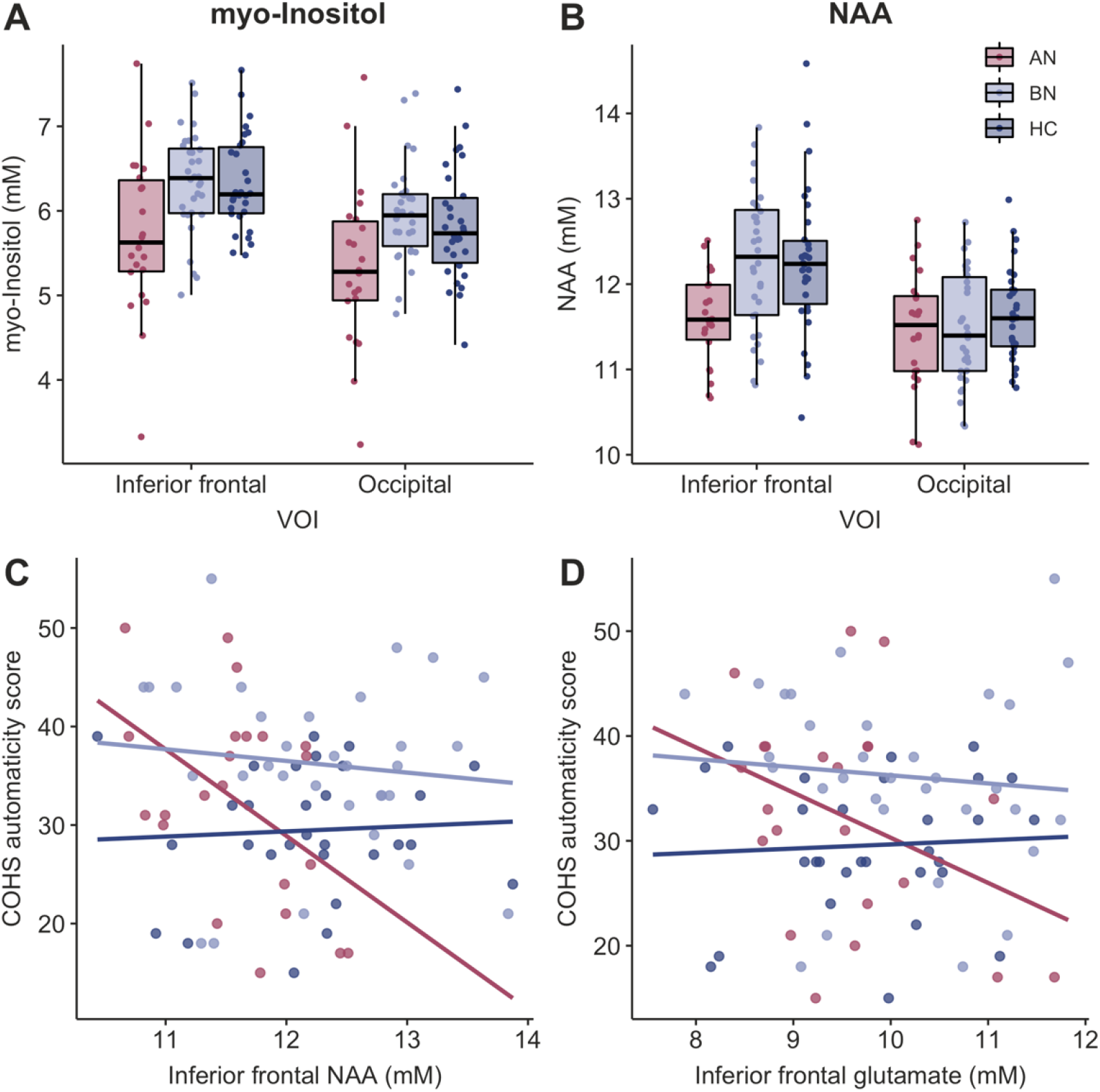
Cerebral metabolites are altered in anorexia nervosa and associated with automaticity. Women with AN-BP, but not BN, had reduced concentrations of **A)** *myo*-inositol (p=.008) and **B)** NAA (p=.007) relative to control participants in right inferior lateral prefrontal and occipital cortices. Metabolite values are corrected for grey matter, white matter and CSF. Exploratory analyses found that associations between right inferior prefrontal metabolite levels and automaticity scores differed between AN-BP and control groups, where reductions in **C)** NAA (p=.021) and **D)** glutamate (p=.039) were related to greater habitual tendencies in women with AN-BP. NAA (F(8, 72)=8.40, p=.02, R^2^_*Adjusted*_=0.12) and glutamate (F(8, 73)=8.36, p=.02, R^2^_*Adjusted*_=0.12) models explained 12% of the variance in automaticity scores.

Sensitivity analyses identified a nominally significant effect of medication status on *myo*-inositol levels, where concentrations were reduced among participants who currently used medication (β(SE)=-0.41(0.18), t(80)=-2.22, p=.03). When controlling for medication use, the observed reduction in *myo*-inositol in AN-BP was nonsignificant (β(SE)=-0.38(0.19), t(80)=-1.97, p=.052); however, we note that the effect estimates in this model overlapped with those of the original model. NAA levels were unrelated to medication status, and NAA remained nominally reduced in women with AN-BP compared to controls with the inclusion of this covariate (β(SE)=-0.35(0.17), t(80)=-2.09, p=.04).

### 3.3 Exploratory analysis of associations between ilPFC metabolites and the habit system

Both AN-BP (β(SE)=6.76(3.08), p=.03) and BN (β(SE)=7.49(2.69), p=.009) groups scored significantly higher than controls on the COHS routine subscale; however, only women with BN reported greater automaticity (β(SE)=6.62(2.15), p=.003). Patient status explained 7% and 8% of the variance in routine (F(2, 81)=4.17, p=.02, R^2^_*Adjusted*_=0.07) and automaticity scores (F(2,81)=4.79, p=.01, R^2^_*Adjusted*_=0.08), respectively. Associations between ilPFC NAA levels and automaticity differed between AN-BP and control groups (group-by-metabolite interaction; β(SE)= -9.75(4.05), p=.019), where lower NAA levels were related to greater automaticity in AN-BP (Figure 3C). Similarly, the relationship between ilPFC glutamate and automaticity was moderated by group such that glutamate was negatively associated with automaticity in AN-BP (β(SE)= -5.75(2.74), p=.039; Figure 3D). These associations remained significant when controlling for medication use (NAA p=.021; glutamate p=.032). All other main and interaction effects between metabolite concentrations, group and habitual biases were nonsignificant.

## 3. Discussion

We used a multi-modal approach to examine neurochemical profiles and goal-directed versus habit systems in women with AN-BP, BN and unaffected controls. Within a highly-controlled inpatient setting, we replicated previous reports of reduced *myo*-inositol and NAA in individuals with AN (15–17), extending these findings to the binge-eating/purging diagnostic subtype. Women with BN did not show significant alterations in the metabolites of interest, which could suggest some diagnostic specificity. Moreover, self-reported habitual tendencies were greater in both patient groups relative to controls while instrumental learning task performance did not differ between groups. We discuss the implications of these results in the following sections.

Decreased *myo-*inositol concentrations in AN-BP could indicate reduced glial and/or myelin integrity. The observation of reduced myo-inositol in both ilPFC and occipital voxels suggests that this difference extends beyond regions that have an established functional role in AN. *Myo*-inositol is considered a marker of glial integrity (50), and it has a key role both in lipid metabolism and myelin sheet structures (51). Whereas demyelination has been associated with increased concentrations of free *myo*-inositol, the specific mechanisms underlying reductions in *myo*-inositol remain unknown. Glial damage or impairment has been suggested to underlie reduced *myo*-inositol in other psychiatric disorders, including schizophrenia and depression (52–54). This mechanism may also generalize to AN as several diffusion-weighted imaging studies have reported increased radial diffusivity in affected individuals (55, 56). Radial diffusivity serves as a measure of cross-fiber diffusion in WM tracts, and greater values indicate dysregulated myelination (57).

However, the combination of reductions in both *myo*-inositol and NAA may be further suggestive of altered axonal-glial signaling in AN-BP. NAA, commonly referenced as a marker of neuronal density, is synthesized from glutamate by mitochondrial enzymes in neuronal cell bodies and axons. The metabolite is then released into the extracellular matrix for uptake by oligodendrocyte cells that, in turn, produce the enzyme, aspartoacylase. Aspartoacylase degrades NAA to aspartate and acetate, and the resulting acetate may serve to maintain myelination (58, 59). NAA therefore seems to have a central role in axonal-glial signaling, where reductions could reflect either reduced neuronal integrity, altered oligodendrocyte functioning or a combination of the two (12). Indeed, *in vivo* structural MRI and postmortem studies report cortical thinning (60, 61) and reduced oligodendrocyte gene expression profiles (62), respectively, in acute AN, lending complementary support to the notion of altered axonal-glial signaling in the illness. Interestingly, as widespread reductions in cortical thickness rapidly normalize with weight restoration (60, 61), cortical pseudoatrophy in AN may partly reflect metabolic disturbances that arise during starvation (63). Our finding of reduced prefrontal and occipital NAA concentrations in AN-BP, but not BN, lends additional support to this theory.

Both patient groups had normal glutamate levels in the ilPFC and occipital cortex; however, ilPFC glutamate was inversely related to automaticity in AN-BP. Disrupted glutamatergic signaling has been associated with automaticity (27) and disinhibition (64) in patients with substance use and neurodegenerative disorders, respectively. Although we provide novel evidence, linking ilPFC glutamate concentrations to habitual biases in AN-BP, this finding was not corrected for multiple comparisons and thus should be interpreted cautiously. Reductions in glutamate or ‘Glx’ have previously been observed in the medial PFC, ACC, occipital cortex and striatum in AN (16, 17), yet variable voxel placement, and assessment of relative versus absolute metabolite concentrations, complicates direct comparison across these studies. Additionally, as prior studies have primarily recruited women with restrictive AN, our results may suggest that reduced glutamate concentrations in AN are specific to the restrictive subtype. We encourage future studies to explore this possibility directly.

We found inconsistent evidence of increased habitual tendencies in women with AN-BP and BN, with notable differences across self-report and task-based measures. Both patient groups reported increased routines relative to the control group; however, automaticity scores were only increased in the BN group. We previously found that women with BN, but not AN-BP, in this cohort have impairments in proactive inhibition (2), and the present findings offer further evidence of distinct self-regulatory deficits across these two disorders. Nevertheless, nonsignificant group differences in slips-of-action task performance highlights outstanding challenges in relation to our understanding of dysregulated learning in eating disorders and its neural correlates. Self-report and task-based measures of related cognitive domains [e.g., self-control (65)] are known to have poor convergent validity, and laboratory-based assessments of goal-directed versus habitual control may be particularly challenging as habits are difficult to induce in humans (66). For example, prior study of patients with acute and recovered AN also failed to identify increased habitual responding on the slips-of-action task (67). Moreover, exploratory analyses linked reduced prefrontal NAA and glutamate with greater automaticity in AN-BP, yet task-based indices of the habit system were unrelated to the cerebral metabolites of interest. This dissociation might suggest that reductions in NAA and glutamate, and associated changes in neuronal integrity and neurotransmission, respectively, are more likely to be associated with habitual traits in AN-BP, as measured by the COHS, rather than more transient and specific habits induced by our task. Since others report deficient goal-directed behavior in AN patients that persists following weight restoration (68), it will be critical to determine if the observed associations between NAA, glutamate and automaticity are specific to acute AN-BP, or if these might reflect a vulnerability to, or persisting consequence of, the illness. Finally, because outcome devaluation tasks are thought to largely index (lack of) goal-directed control (66), alternative approaches that move beyond simplistic dichotomies in cognitive (neuro)science will be central to refining knowledge of the processes underlying seemingly habitual behavior [reviewed by (24)].

Although this work has notable strengths, some limitations should be considered. First, approximately 30% and 45% of women with BN and AN-BP, respectively, were taking prescribed psychotropic medication, which may affect glutamate turnover and glucose metabolism (69). When controlling for current medication use in sensitivity analyses, we identified similar reductions in *myo*-inositol and NAA levels in the AN-BP group compared to controls, albeit at nominal and trend-level significance, respectively. This would suggest that our findings in AN-BP are at least partially robust to medication use. However, as medication use likely covaries with other factors (e.g., illness severity, treatment access), further work is needed to precisely quantify the effects of psychotropic medication on ^1^H-MRS spectra, particularly in patients with chronic psychiatric illness (70). Second, the use of single-voxel ^1^H-MRS precluded assessment of the role of metabolites in brain activation or structure due to low spatial resolution. Alternative techniques, such as multi-voxel functional spectroscopy [e.g., (71)], would facilitate analysis of temporal associations between metabolite concentrations, functional activity and cortical morphometry. Third, we were unable to quantify other neurotransmitters, such as GABA, that may have functional relevance to eating disorders but cannot be reliably estimated at 3T. As such, future ultra-high-field ^1^H-MRS studies of eating disorders are warranted.

In summary, these findings lend novel insight into neural chemistry of AN-BP and BN, suggesting that these illnesses show distinct neurobiological profiles despite shared symptomatology. Our results support the role of the right ilPFC in habitual behavior, specifically in AN-BP, yet assessment of habit learning remains challenging due to relatively poor convergent validity across measures. Future studies should integrate ^1^H-MRS and task-based functional neuroimaging to further elucidate associations between cerebral metabolites and cognition in eating disorders.

## Data Availability

Data will be made available upon reasonable request following manuscript publication. Interested individuals may contact Dr Margaret Westwater.

## Data Availability

All data related to this work is available upon request

## Acknowledgements

The authors thank the participants for their time and dedication to the study; the nursing staff and radiographers for assisting the study protocol; and Dr. Dinesh Deelchand for sharing code for the MRS voxel segmentation. They also wish to thank the Beat eating disorder charity for advertising the study. Funding was provided by the Bernard Wolfe Health Neuroscience Fund to PCF and HZ and a Wellcome Trust Investigator Award to PCF (Reference No. 206368/Z/17/Z). MLW was supported through the NIH-Oxford-Cambridge Scholars Program and a Cambridge Trust fellowship. AGM was supported by a Holt Fellowship. The Wellcome Trust/NIHR Clinical and Translational Research Facilities and the Wolfson Brain Imaging Centre provided equipment and support staff for the study. This research was supported by the NIHR Cambridge Biomedical Research Centre (BRC-1215-20014). The views expressed are those of the author(s) and not necessarily those of the NIHR or the Department of Health and Social Care.

## Notes

### Competing Interest Statement

The authors have declared no competing interest.

### Author Declarations

The Cambridge East Research Ethics Committee approved the study (Ref. 17/EE/0304), and all procedures were performed in accordance with local regulations.

