## Supplementary Table 1 for "Characterizing cerebral metabolite profiles in anorexia and bulimia nervosa and their associations with habitual behavior"

1. **Methods**

**Data analysis – Behavior**

Performance on the instrumental learning task [accuracy, reaction time (RT)] was evaluated using linear-mixed effects models (LMMs) and multiple regression. Outlier RT values were defined as those >1.5 times the interquartile range above the 75^th^ percentile and below the 25^th^ percentile, and for the instrumental learning stage, outliers were evaluated for each task block. A rank-based inverse normal transformation was applied to the remaining RTs to minimize positive skew when necessary [R package ‘RNOmni’ (1)].

Each model included fixed effects of group (AN-BP>HC, BN>HC) and task condition (block for the instrumental learning stage; stimulus or outcome value for discrimination and slips-of-action stages, respectively), where random intercepts for task condition were nested within the random effect of the subject. Group differences were evaluated using non-orthogonal contrast coding, in which AN-BP and BN groups were compared to controls (e.g., treatment contrasts). Finally, multiple regression was used to assess group differences in sensitivity to outcome devaluation (stage 2) and mean differences in the proportion of responses to valued and devalued outcomes.

**Data analysis – Neuroimaging**

***Data acquisition and pre-processing***

Spectra were collected using a semiLASER (sLASER) sequence, which was selected over a vendor-provided PRESS protocol due to its improved localization, spectral quality and replicability (2, 3). Specifically, previous sLASER experiments have obtained highly reproducible neurochemical profiles for five major metabolites, including the three metabolites of interest in the present study (i.e., glutamate, NAA, *myo*-inositol), where coefficients of variance were <5% at both 3 and 7 Tesla (4).

Pre-processing of ^1^H-MRS spectra was performed using the MRSpa MATLAB package (<https://www.cmrr.umn.edu/downloads/mrspa/>). Raw MRS free induction decays (FIDs) were corrected for eddy current effects, and a zero-order phase correction was applied. FIDs were corrected for temporal (or “frequency”) and phase drifts using the cross-correlation method, which minimizes the frequency and phase difference, respectively, between single-shot MRS data. Correction of frequency and phase drifts served to mitigate broadening of the spectra, improve the signal-to-noise ratio and prevent line shape distortion (5). Finally, FIDs were then visually inspected for outlier transients (e.g., those containing a large residual water peak or noise-only spectrum) as these would be indicative of large motion artifacts. Outlier FIDs (<10%) were removed from 26 ilPFC and 18 occipital spectra before summation.

***Voxel segmentation***

To control for partial volume effects on metabolic estimation, we calculated the proportion of grey matter (GM), white matter (WM) and cerebrospinal fluid (CSF) in each voxel of interest (VOI). Whole-brain, voxel-based tissue segmentation of the T1-weighted anatomical scan was conducted in SPM12 (Wellcome Department of Clinical Neurology, London), using ICBM Tissue Probabilistic Atlases. Each VOI was then coregistered to the anatomical image, and the percentage of GM, WM and CSF tissue within the VOIs was computed using both the GM and WM probability maps, using custom Matlab code provided by Dr Dinesh Deelchand of the University of Minnesota.

2. **Results**

**Behavior - Explicit knowledge of stimulus, response and outcome associations**

Upon completion of the instrumental learning task, participants responded to a set of questions that indexed their explicit knowledge of stimulus, response and outcome associations. Questionnaire data were available from 80 participants (n=20 AN-BP, n=32 BN, n=28 HC), and knowledge of stimulus-outcome pairings and correct responses did not differ between groups (all p’s > .05; see Supplemental Figure 1).

**Table S1**. Spectral quality metrics

| **Characteristic** | AN  M(SD) | BN  M(SD) | HC  M(SD) | *F* statistic | *P*-value |
| --- | --- | --- | --- | --- | --- |
| Water line width (Hz) |  |  |  |  |  |
| *Right inferior frontal cortex* | 8.35 (0.58) | 8.54 (0.78) | 8.56 (0.83) | 0.56 | .57 |
| *Right occipital cortex* | 7.63 (0.31) | 7.76 (0.45) | 7.82 (0.44) | 1.25 | .29 |
| Sign-to-noise-ratio |  |  |  |  |  |
| *Right inferior frontal cortex* | 49.73 (7.23) | 50.66 (7.43) | 51.7 (8.32) | 0.42 | .66 |
| *Right occipital cortex* | 51.64 (11.84) | 50.78 (11.71) | 54.60 (12.58) | 0.83 | .44 |
| Glutamate CRLB |  |  |  |  |  |
| *Right inferior frontal cortex* | 4.22 (0.75) | 4.22 (0.79) | 4.23 (0.86) | .003 | .99 |
| *Right occipital cortex* | 5.64 (2.34) | 5.45 (2.01) | 5.00 (1.20) | 0.83 | .44 |
| Myo-inositol CRLB |  |  |  |  |  |
| *Right inferior frontal cortex* | 4.32 (1.09) | 4.03 (0.78) | 3.80 (0.61) | 2.53 | .09 |
| *Right occipital cortex* | 4.32 (1.36) | 4.10 (1.25) | 3.97 (0.96) | 0.56 | .57 |
| NAA CRLB |  |  |  |  |  |
| *Right inferior frontal cortex* | 2.23 (0.43) | 2.13 (0.42) | 2.00 (0.37) | 2.04 | .14 |
| *Right occipital cortex* | 2.32 (0.89) | 2.52 (0.89) | 2.30 (0.53) | 0.70 | .50 |
| % Grey matter |  |  |  |  |  |
| *Right inferior frontal cortex* | 56.65 (7.33) | 56.30 (7.35) | 54.21 (6.47) | 0.99 | .38 |
| *Right occipital cortex* | 42.91 (4.37) | 43.08 (5.54) | 43.28 (4.48) | 0.04 | .96 |
| % White matter |  |  |  |  |  |
| *Right inferior frontal cortex* | 35.37 (8.86) | 35.02 (9.20) | 38.19 (7.62) | 1.21 | .30 |
| *Right occipital cortex* | 52.80 (5.01) | 52.92 (6.85) | 53.14 (4.82) | 0.02 | .97 |
| % CSF |  |  |  |  |  |
| *Right inferior frontal cortex* | 7.19 (2.17) | 7.82 (3.08) | 6.82 (2.18) | 1.20 | .31 |
| *Right occipital cortex* | 3.94 (1.81) | 3.63 (2.02) | 3.21 (1.53) | 1.10 | .34 |

**Notes:** CRLB = Cramer-Rao lower bound, NAA = *N-*acetyl aspartate, CSF = cerebrospinal fluid.

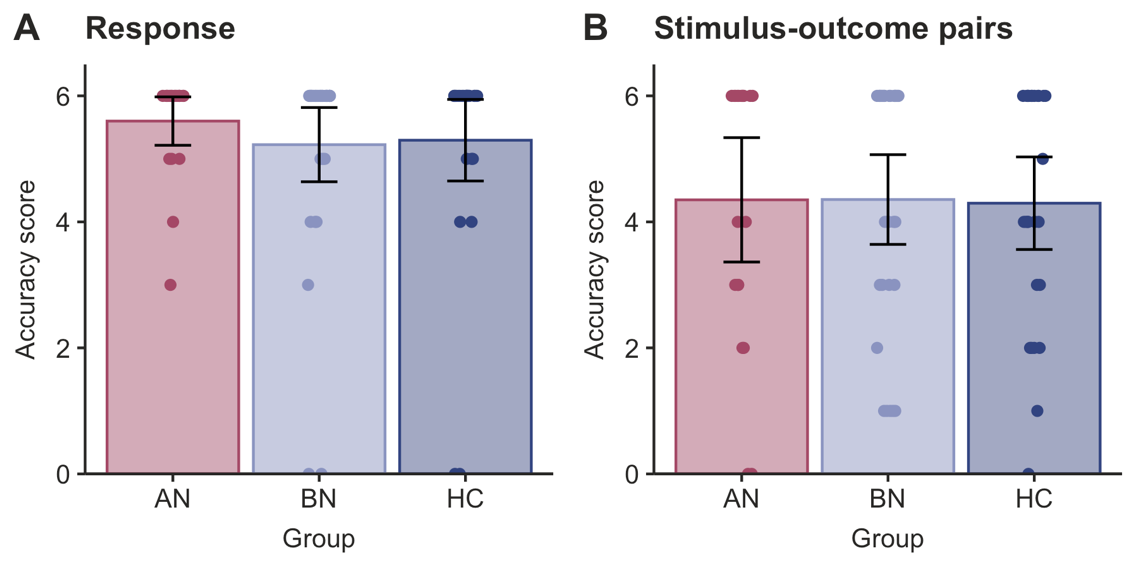

**Figure S1.** *Explicit knowledge of stimulus, response and outcome associations.* AN-BP, BN and control groups did not differ significantly in their explicit knowledge of stimulus contingencies or responses. Error bars = 95% CI.
